# Importation risk and preparedness priorities across Africa in the 2026 Bundibugyo Ebola outbreak

**DOI:** 10.64898/2026.07.21.26358330

**Authors:** Federico Fanelli, Francesco Parino, Maria Amalia Pelle, Boxuan Wang, Yap Boum, Mosoka Fallah, Placide Mbala, Reagan Luvande Okingo, Vincent Ronin, Yazdan Yazdanpanah, Chiara Poletto, Eugenio Valdano, Vittoria Colizza

## Abstract

The ongoing 2026 Bundibugyo Ebola outbreak in the Democratic Republic of the Congo raises concerns about regional dissemination across Africa. Anticipating where imported infections are most likely to occur, and whether recipient countries are prepared to detect and contain them, is essential for prioritizing surveillance and response. We developed an integrated spatial framework combining high-resolution epidemic and population data, air and land mobility, geographical accessibility, conflict-adjusted travel times to estimate the relative risk of Ebola importation across African countries. We contextualized importation risk using different national readiness indicators. We quantified overall, air-mediated, and land-mediated importation risk under the current epidemic situation and evaluated outbreak amplification scenarios in major regional hubs. Importation risk was highly concentrated geographically, with neighbouring countries accounting for 93% of the estimated continental risk. However, high-risk destinations extended beyond the immediate neighbours, whereas some neighbouring countries remained at low importation risk, reflecting the complementary roles of land and air mobility in shaping regional dissemination. South Sudan, Rwanda, Zambia, Kenya, Ethiopia, Tanzania, Burundi, Angola, South Africa, the Central African Republic, and Nigeria were estimated at high importation risk, but differed markedly in healthcare system capacity and emergency preparedness and response. Readiness gaps were largest in South Sudan, Burundi, and the Central African Republic. Botswana, Burkina Faso, Cameroon, Chad, and Ghana emerged as a second tier of preparedness priorities, combining intermediate-high importation risk with low readiness. Under outbreak amplification scenarios, the highest-risk countries remained largely stable, but additional countries entered the high-risk group and continental importation pressure increased substantially, highlighting the need for dynamic preparedness planning. Integrating high-resolution epidemiological data with complementary land and air mobility provides a refined geography of regional importation risk and preparedness across Africa, supporting better targeted surveillance, preparedness investments, and international support while allowing priorities to be rapidly updated as outbreaks evolve.

## INTRODUCTION

The ongoing Bundibugyo Ebola virus disease outbreak in eastern Democratic Republic of the Congo (DRC) has substantial potential for regional spread. Importations into other African countries, particularly where preparedness and response capacities are limited, could turn the outbreak into a broader public health emergency. As of 22 August 2026, the outbreak - previously declared a Public Health Emergency of International Concern (PHEIC)^1^ - has reached 5,514 confirmed cases across the provinces of Ituri, North Kivu, South Kivu, Haut-Uélé, Bas-Uélé, and Tshopo in eastern DRC^2^. The spread to neighboring Uganda resulted in 20 confirmed cases, before the Africa Centres for Disease Control and Prevention (Africa CDC) and the World Health Organization declared the epidemic over on August 27, 2026, following 42 consecutive days without a new confirmed case^3^. Several factors increase the risk of further dissemination. The virus is spreading in both urban centers and remote rural settings, where response efforts face distinct operational challenges^4,5^. Ongoing armed conflicts hamper surveillance and healthcare delivery, while the outbreak epicenter lies close to highly porous international borders characterised by intense movements driven by trade, mining, and population displacement^4,5^. There, informal crossings outside official points of entry further complicate surveillance and compromise border control measures^5^.

Regional spread is facilitated by the combination of prolonged incubation, extensive cross-border mobility, and, potentially, air transportation. During the incubation period, infected individuals may travel undetected, seeding infections into neighbouring countries or more distant destinations, as shown by the case reported in France on June 24^3^. Evidence from past Ebola epidemics also shows that international dissemination can occur despite border screening and reductions in travel^6,7^. During the 2014–2016 West Africa epidemic, imported cases were reported in seven countries outside the source region^7^, including three in Africa. Additional international exportations may therefore occur if the epidemic expands.

Preventing regional amplification requires not only rapid control at the outbreak source but also preparedness in receiving countries to detect and contain imported infections before sustained local transmission becomes established. In the absence of licensed vaccines and specific treatments against Bundibugyo virus, early detection and implementation of core public health measures remain the cornerstone of outbreak control^8^.

Anticipating where importations are most likely to occur remains challenging. Previous modeling studies have estimated the regional^9,10^ and international^11–13^ spread of Ebola Bundibugyo, but have simplified mobility within Africa by relying on air mobility flows or restricting the analysis to a limited set of neighboring countries. Regional dissemination across the continent, however, is dominated by complex patterns of air and land mobility, and heterogeneous geographical accessibility, which are not adequately captured by air mobility alone. Moreover, despite the 2018–2020 DRC Ebola outbreak (also declared a PHEIC) occurring in the same North Kivu and Ituri provinces, the present outbreak is distinct in its rapid spread and geographical extent, making it difficult to rely on past evidence for risk planning. At the same time, African countries differ substantially in healthcare capacity, surveillance systems, available medical supplies, emergency preparedness and operational response capabilities, as well as risk communication^5,14,15^. Integrating these complementary dimensions is essential for actionable preparedness.

We developed an integrated analytical framework to estimate the importation risk of Ebola Bundibugyo in African countries in relation to national readiness, by combining epidemiological and high-resolution spatial population data, commercial air transportation, land mobility, geographical accessibility, conflict-adjusted travel times, and different readiness scores. We further investigated how the geography of importation risk would change under different epidemic amplification scenarios. By contextualizing importation risk with national readiness, our framework provides operational evidence to support regional preparedness, surveillance prioritization, and international coordination for the current outbreak.

## METHODS

We assessed the importation risk for regional spread of Ebola virus disease for the 2026 Bundibugyo outbreak in eastern DRC using a high-resolution multi-layer framework integrating epidemiological, mobility, population, accessibility, capacity, and response data (**Supplementary Note 1, Section S0**). At the time of analysis, ongoing transmission was reported only in DRC; the earlier outbreak in Uganda following case importation from DRC had been declared over^16^. We therefore considered affected areas in DRC as the only sources of infection. Uganda was not considered as a potential destination, as its previous importation and subsequent outbreak response placed it in a different epidemiological and preparedness context from countries not yet affected by the outbreak. The analysis focused on the affected health districts in DRC and evaluated the epidemic activity of these areas and their connectivity to the rest of Africa through both air and land mobility flows, accounting for travel time, conflicts, length of incubation period, and proximity to transportation hubs (**Fig. 1**).

**Figure 1.**
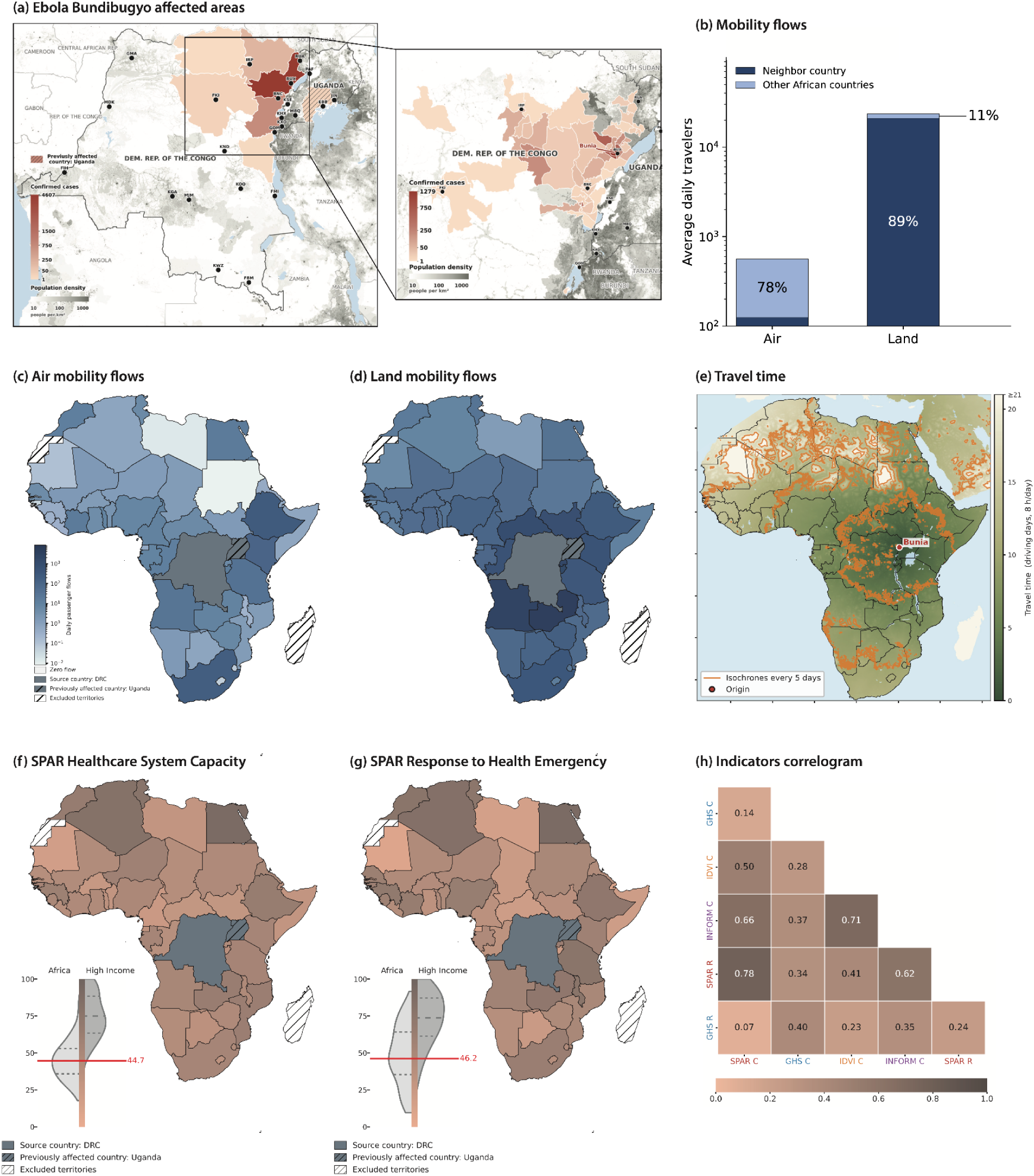
Epidemiological context, mobility flows, travel time, and preparedness indicators. **(a)** Spatial distribution of reported Bundibugyo Ebola-affected areas in the Democratic Republic of the Congo, with the inset showing the affected area in greater detail. The color code indicates the cumulative number of confirmed cases per affected health district (DRC) and ADM3 unit (Uganda), overlaid on population density (grey). Uganda, which was previously affected following case importation from DRC but was declared Ebola-free on 27 August, is shown with hatching. **(b)** Average daily outbound mobility flows from the DRC, stratified by travel mode and by neighboring vs. other African countries. **(c, d)** Average daily air (c) and land (d) mobility flows from the DRC to African countries, on a logarithmic colour scale. DRC, the source country, is shown in grey. Uganda, being declared Ebola-free, is excluded from the flow analysis and is shown in grey with black hatching. Excluded territories are shown in white with black hatching, while countries with no estimated flow are indicated separately. **(e)** Estimated overland travel time from the outbreak source expressed in driving days assuming 8 hours of travel per day; isochrones are shown for every five days. The map shows the example of travel time from Bunia, the health district with the largest activity as of 22 August 2026. Travel time was computed from all affected health districts and ADM3 in DRC. **(f, g)** Country-level readiness scores based on SPAR healthcare system capacity (f) and SPAR preparedness and response to health emergencies (g); the inset violin plots compare the distribution of indicator values across African and high-income countries, and the red line marks the median across African countries. DRC, the source country, is shown in grey. Uganda, being declared Ebola-free, is excluded from the analysis and is shown in grey with black hatching. Excluded territories are shown in white with black hatching, while countries with no estimated flow are indicated separately. **(h)** Correlation matrix of selected healthcare system capacity (C) and emergency response (R) indicators derived from the SPAR, GHS, IDVI, and INFORM frameworks, computed across the African countries; the corresponding principal component analysis is reported in **Supplementary Note 1**.

Cumulative confirmed cases were obtained from the Institut National de Santé Publique (INSP) of the DRC in the daily situation report number 100 published on 23 August 2026^2^(**Fig. 1a**). To account for geographically heterogeneous reporting rates, we used the proportion of alerts verified among the alerts received in each DRC province, averaged over the two weeks preceding 23 August 2026^2^ (**Supplementary Note 1**). We explored alternative values in the sensitivity analysis. Population data were obtained from the 2024 health-zone population projections produced by the DRC Information Management Working Group and disseminated by the United Nations Office for the Coordination of Humanitarian Affairs (OCHA) through the Humanitarian Data Exchange Platform^17^. These datasets were used to compute cumulative attack rates in affected areas and to account for differences in local epidemic intensity across source health districts when estimating their relative contribution to international exportation.

International mobility in Africa was characterized through complementary air and land mobility. Air mobility was derived from the International Air Transport Association (IATA) data on origin–destination passenger flows between any pair of commercial airports and accounting for connections at intermediate airports (**Fig. 1c**). Land mobility was then obtained as residual mobility by subtracting air mobility from the Global Transnational Mobility Dataset 2.0^18^ (GTMD2), which provides estimates of overall population movements between countries (**Fig. 1d**). Mobility volumes spanned more than five orders of magnitude across country pairs, ranging from less than a traveller per day to more than 10^4^ daily travellers on the most connected routes. Land mobility dominated regional connectivity around the outbreak area, accounting for 89% of cross-border mobility with neighbouring countries. The contribution of air transportation was substantially smaller (by two orders of magnitude) but geographically broader, with about three quarters of movements linking the source area to African countries beyond its immediate neighbors (**Fig. 1b**).

To characterize spatial accessibility from the outbreak area and the potential for land-based spread within the incubation period, we computed travel-time surfaces using a global motorized-travel friction surface map^19^ integrating road networks, land cover, rivers, topography, and transport infrastructure. Travel times were estimated from each affected health district in DRC to all destination countries in Africa. As armed conflict can substantially alter travel time, our estimates were adjusted for conflict-related disruptions using the most recent conflict data at Administrative Level 2 (ADM2) obtained from the Armed Conflict Location & Event Data Project^20^ (ACLED), and calibrated against Google Maps driving-time estimates (**Supplementary Note 1**). The conflict-adjusted travel-time estimates were independently validated against Google Maps driving times for 10 out-of-sample trips between African capitals, not included in the model fitting, and showed better agreement with observed travel times than friction-based estimates not accounting for conflict (**Supplementary Note 1**). **Fig. 1e** shows as an example the estimated travel time from Bunia, the health district reporting the largest epidemic activity, under the assumption of 8h driving time per day; additional daily driving times were considered for sensitivity.

Populations in DRC health districts were linked to land and air mobility networks through spatial allocation models (**Supplementary Note 1**). For land travel, the share of the population in each administrative unit travelling to each destination country was estimated through a gravity model calibrated on observed land mobility patterns. Uncertainty in the fitted gravity parameters was assessed using an origin-country cluster bootstrap and propagated through the land-mobility allocation and importation-risk calculation (**Supplementary Note 1**). For air travel, it was assigned using weights proportional to population size and decreasing with the distance to each airport of the country. The same spatial allocation framework was also used in the amplification scenarios (see below).

Border restrictions and closures related to the current outbreak were not considered, because the implemented measures were partial, temporary and largely limited to official points of entry. Also, available data on cross-country mobility suggested no meaningful reduction in cross-border movement occurred during such restrictions (**Supplementary Note 1**).

Throughout this study, we used the term importation risk to denote a relative measure, defined as the probability that an infected individual departing from the affected areas reaches a given African destination country, conditional on initiating travel (**Supplementary Note 2**). A country’s importation risk represents its share of the modelled infected-traveller flow from the affected areas across unaffected African countries. As such, importation risk sums to one across destination countries.

Importation risk for each African country was estimated from air and land mobility, both separately to quantify air- and land-mediated risk and jointly to obtain the overall importation risk (**Supplementary Note 2**). We assumed that Ebola infected cases could travel only in their latent phase, as symptomatic patients would be expected to either remain in the affected country for clinical management or, if transferred internationally, travel under medically supervised and controlled conditions. Therefore, for land mobility, travel times were explicitly incorporated to estimate the destination reached before the onset of symptoms. For air travel, journey times were considered negligible relative to the incubation period. Importation risks were weighted by the cumulative attack rate in each affected administrative unit to account for geographically heterogeneous epidemic activity. The framework did not distinguish travellers by their country of residence; importation risk was estimated from the movement of infected individuals departing from affected areas, irrespective of whether infection occurred among local residents or visitors. Overall importation risk was estimated by combining air and land mobility flows, while air- and land-mediated risks were estimated using the corresponding mobility flows separately. For each of these three estimates, importation risk values were independently normalized across destination countries to sum to 1. Importation risk was further stratified by border distance (i.e., minimum number of border crossings) from DRC.

National epidemic readiness was assessed using several complementary frameworks, including the World Health Organization (WHO) State Parties Self-Assessment Annual Reporting^15^ (SPAR, 2025), the Global Health Security Index^21^ (GHS, 2021), the Infectious Disease Vulnerability Index^22^ (IDVI, 2016), and the Epidemic Risk Index^23^ (INFORM-WHO, 2020). From each framework, we extracted indicators describing *Healthcare system capacity* and *Emergency preparedness and response* (**Supplementary Note 1**). Correlation and principal component analyses were performed to assess agreement and complementarity across indicators (**Fig. 1h**; **Supplementary Note 1**). Given its complete coverage of African countries, recent update, and direct relevance to the current WHO Framework for Health Emergency Preparedness and Response Capabilities for national public health agencies^24^, SPAR was selected as the primary epidemic readiness metric for the main analysis (**Fig. 1fg**). Corresponding GHS indicators, complementary to SPAR indicators, were used in sensitivity analyses. All values are defined on a 0-100 scale, with 100 corresponding to the highest capacity or response value.

All quantitative analyses were performed using the continuous importation risk and readiness measures; continuous values are reported in the GitHub repository. Countries were then classified into quartiles of importation risk, healthcare system capacity, and emergency preparedness and response, corresponding to high (n=11), intermediate-high (n=12), intermediate-low (n=12), and low categories (n=11), to facilitate interpretation and discussion of preparedness priorities.

Amplification scenarios were designed as plausible stress-test scenarios^11^ to assess how geographical expansion of the epidemic beyond the currently affected area into major regional hubs could modify importation risk across Africa. Tested locations included Kinshasa, the capital of the currently affected country (DRC); Kampala (Uganda), a major regional hub in a country previously affected during the outbreak; and the capitals of neighboring countries scoring highest in importation risk: Juba (South Sudan), Kigali (Rwanda), Bujumbura (Burundi), Nairobi (Kenya), and Lusaka (Zambia). These cities were selected because of their strategic role as major population and transportation hubs in high-risk countries, allowing us to evaluate how epidemic expansion into highly connected urban centres could reshape preparedness priorities. They differ in population size, geographical location, and transportation connectivity. Bujumbura was selected instead of Gitega as it is the economic capital and largest city of Burundi. In each scenario, the simulated epidemic activity in the selected city was added to the observed epidemic activity in the currently affected areas in DRC, representing a multi-focal outbreak configuration. A cumulative attack rate of 5 cases per 100,000 persons was assumed in each city, consistent with the median cumulative attack rate reported across affected health zones (7.4 cases per 100,000 persons). Sensitivity analyses using 1 and 10 cases per 100,000 persons were performed to assess the robustness of the results to the assumed level of epidemic activity. To quantify changes in the magnitude of importation under the amplification scenarios, we calculated the fold change in importation pressure relative to the current epidemic situation (**Supplementary Note 2**). For each destination country, this quantity was defined as the ratio of the mobility flow weighted by the epidemic activity reaching that country under the amplification scenario to the corresponding flow under the current epidemic situation. This measure indicates how many times greater the modelled inflow of infected travellers would be under an amplification scenario.

## RESULTS

Overall importation risk was concentrated geographically (**Fig. 2a**). The 11 countries in the high-risk quartile – South Sudan, Rwanda, Zambia, Kenya, Ethiopia, Tanzania, Burundi, Angola, South Africa, Central African Republic, and Nigeria – accounted for 93% of the total estimated importation risk (**GitHub Repository**), and formed a largely contiguous regional belt surrounding the affected area, with the exception of Kenya, Ethiopia, South Africa, and Nigeria, which were geographically more distant (at border distance 2, 2, 3, and 3 from the outbreak source, respectively). All countries directly bordering DRC were classified as high risk, except the Republic of the Congo at low risk. Neighboring countries alone accounted for 73% of the total estimated risk, while countries located two border crossings away contributed an additional 21%, highlighting the strong spatial concentration of potential spread around the outbreak source (**Fig. 2f**).

**Figure 2.**
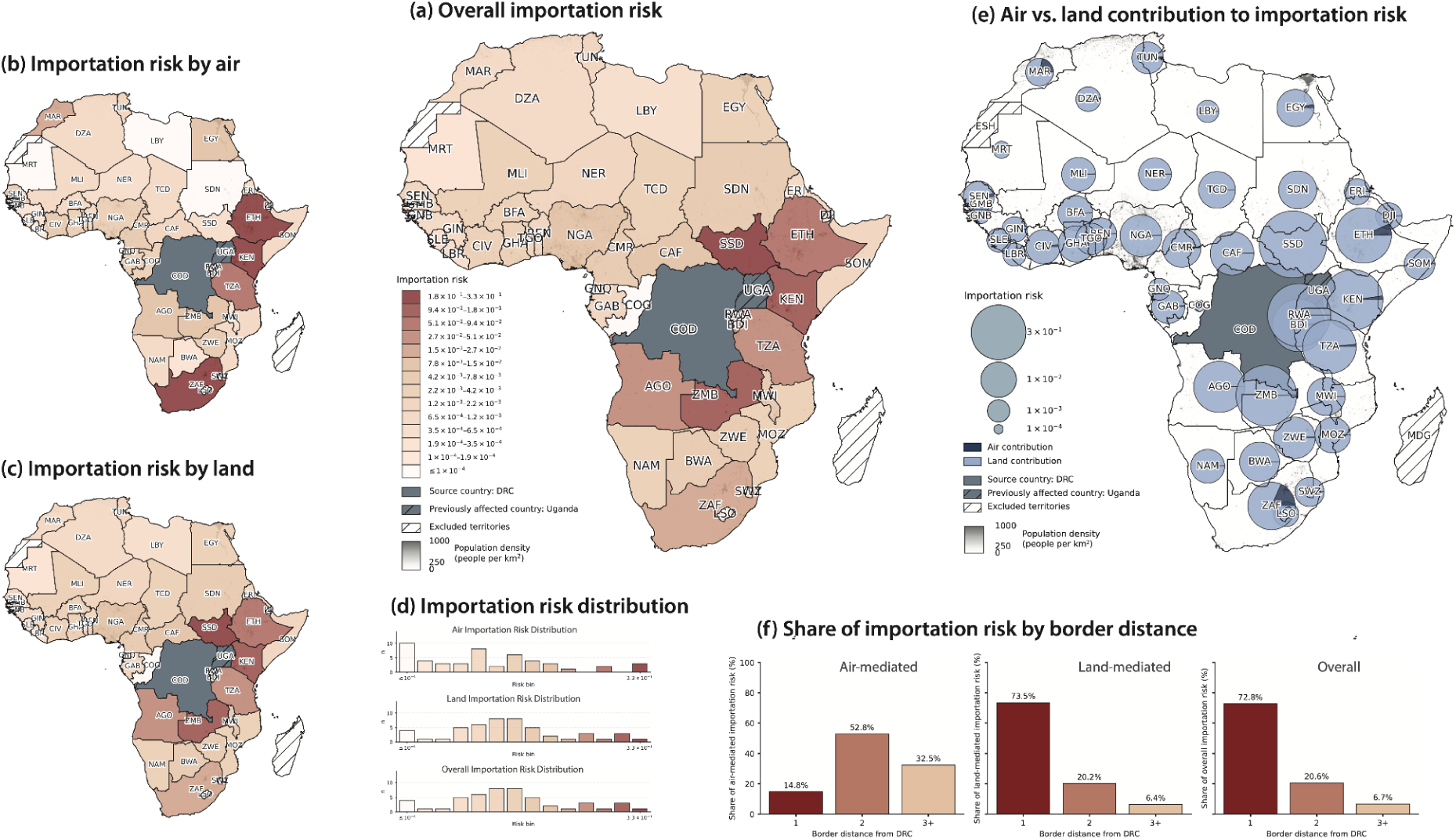
Importation risk of Bundibugyo Ebola across African destination countries under the current epidemic spread. **(a)** Overall importation risk from the Democratic Republic of the Congo (DRC) to each African country, coloured on a categorical logarithmic scale, with population density shown in grey in the background. **(b, c)** Importation risk from the DRC to each African country decomposes into (b) air-mediated and (c) land-mediated risk, shown on the same scale as panel (a). **(d)** Distribution of importation risk considering air-mediated risk (top), land-mediated risk (center), and overall risk (bottom). The y-axis gives the number of countries (n) in each bin. **(e)** Overall importation risk (coded by the circle size) per country along with its relative contribution of air and land mobility pathways (pie shares with dark blue indicating air mobility and light blue indicating land mobility). Population density is shown in grey in the background. **(f)** Share of importation risk by border distance from the source countries, shown separately for the air-mediated risk, land-mediated risk, and overall risk. Border distance is the smallest number of border crossings from DRC to the destination country. In all maps, DRC, the source country, is shown in grey; Uganda, being declared Ebola-free, is excluded from the analysis and is shown in grey with black hatching; excluded territories are shown in white with black hatching, while countries with no estimated flow are indicated separately.

The observed importation risk pattern was primarily driven by land mobility (**Fig. 2b,c,e**). Risk rankings based on overall importation risk largely mirrored those obtained using land mobility alone (**Supplementary Note 1**). As land-mediated importation risk decreased gradually with border distance, countries at highest overall risk were also those most strongly connected to the affected area through land mobility. Air mobility instead generated a different importation risk distribution (**Fig.2d**) and a partially distinct risk geography. Air-mediated importation risk remained relatively high within three border crossings of the outbreak source and decreased sharply at larger distances, with South Africa and Nigeria representing notable distant exceptions (**Supplementary Note 1**). Kenya and Ethiopia ranked among the five countries at highest risk of importation mediated by each mobility layer (**Fig. 2b,c**; **Supplementary Note 1**), reflecting their strong regional connectivity, despite both being located two border crossings from the outbreak source. Kenya, Ethiopia, South Africa, and Nigeria were the only non-neighbouring countries classified as high risk overall, but showed different contributions of air and land mobility. Kenya ranked similarly for air and land importation risk (third and fourth, respectively); Ethiopia and South Africa ranked substantially higher for air than for land importation risk (first vs fifth and second vs ninth, respectively); while Nigeria showed the opposite pattern (16th for air-mediated importation risk vs 11th by land) (**Supplementary Note 1**). Country-level importation-risk rankings were highly robust to uncertainty in the fitted gravity parameters (median Spearman correlation with the baseline ranking: 0.9999, p-value< 0.001; **Supplementary Note 1)**.

National epidemic readiness varied considerably across Africa and showed no significant association with importation risk (**Fig. 3**; Spearman r=0.12, p-value=0.42 with healthcare system capacity, Spearman r=0.24, p-value=0.11 with emergency preparedness and response). Countries with similar levels of exposure displayed markedly different healthcare system capacity and emergency preparedness and response scores (**Fig. 3a,b**). Among the 11 high-risk countries, three (Zambia, Ethiopia, and Tanzania) combined high healthcare system capacity with high emergency preparedness and response, while Nigeria exhibited intermediate-high healthcare system capacity and high emergency preparedness and response (**Fig. 3c**). In contrast, South Sudan, Burundi, and the Central African Republic had high importation risk but low epidemic readiness. South Sudan exhibited low healthcare system capacity and intermediate-low emergency preparedness and response; Burundi exhibited intermediate-low healthcare system capacity and low emergency preparedness and response; the Central African Republic ranked low on both indicators. The specific preparedness gaps underlying these low readiness levels differed across countries: South Sudan showed particularly low human resources and infection prevention and control (IPC) capacity, together with limited capacity at points of entry; Burundi showed gaps in laboratory capacity, human resources, risk communication and community engagement (RCCE), and points of entry; and the Central African Republic showed particularly low IPC, laboratory and human resource capacity, together with gaps in RCCE and points of entry (**Supplementary Tables S12 and S13**). The remaining high-risk countries (Rwanda, Kenya, Angola, and South Africa) showed intermediate levels of readiness.

**Figure 3.**
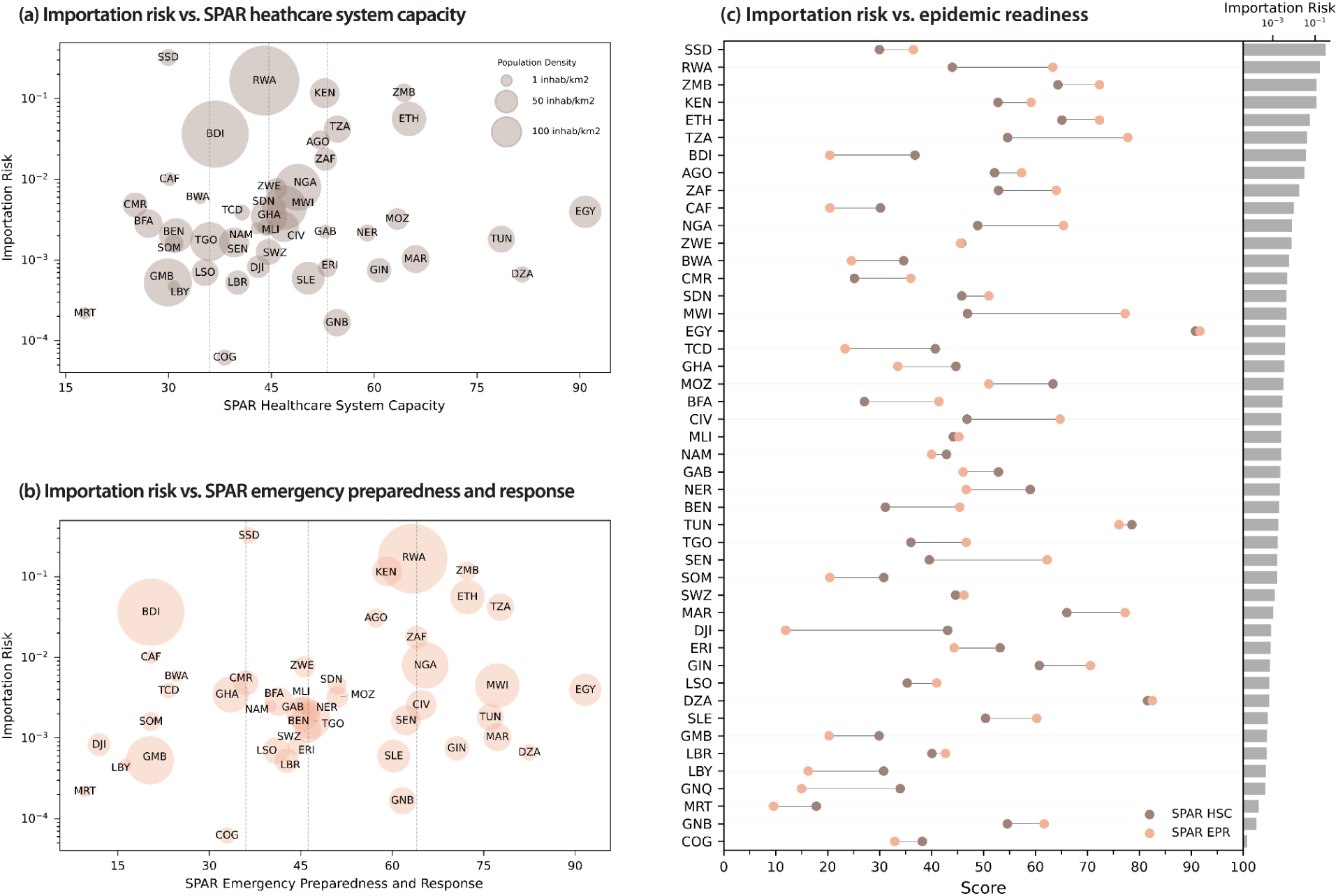
Importation risk as a function of country readiness. **(a, b)** Scatter plots of overall importation risk versus SPAR Healthcare System Capacity (a) and SPAR Emergency Preparedness and Response (b) scores. The y-axis is displayed on a logarithmic scale. Circle size is proportional to the population density of each country. The three vertical dashed grey lines indicate the 25th, 50th (median), and 75th percentiles of the corresponding SPAR epidemic readiness dimension. **(c)** SPAR Healthcare System Capacity and SPAR Emergency Preparedness and Response scores for African countries ranked by overall importation risk (from highest risk at the top, to lowest risk at the bottom). The corresponding importation risk values are shown in the inset bar plot on the right, whose risk axis is displayed on a logarithmic scale.

Similarly, countries with intermediate-high importation risk had variable levels of readiness. Four countries exhibited high scores of readiness, in either the healthcare system capacity (Mozambique), or the emergency preparedness and response (Malawi, Côte d’Ivoire), or both (Egypt). Botswana instead had low scores in both indicators, while Burkina Faso and Cameroon showed low healthcare system capacity (together with intermediate-low response), and Chad and Ghana showed low emergency preparedness and response (together with intermediate-low capacity). Among these countries, Botswana main gaps concerned health emergency management, human resources and IPC, while Chad showed particularly low RCCE and point-of-entry capacity, and Ghana showed particularly low health emergency management capacity (**Supplementary Tables S12 and S13**).

We next examined how importation risk would change if sustained local transmission became established in major regional hubs of countries currently at high risk (**Fig. 4**). Across the seven scenarios tested, the set of 11 high-risk countries remained largely unchanged and closely matched that estimated under the current epidemic spread. Specifically, South Sudan, Rwanda, Zambia, and Kenya remained among the 5 countries at highest risk of importation (**Fig. 4a**). Only a few countries entered the high-risk group in some of the amplification scenarios, as was the case of the Republic of the Congo under the expansion of the Ebola epidemic in Kinshasa. In contrast, the fold change in importation pressure relative to the current epidemic situation varied substantially across scenarios (**Fig. 4b**). Epidemic amplification in Nairobi and Lusaka produced the largest increase in continental importation (1.5-fold increase), and especially in southern Africa (up to 4-fold). Amplification in Kampala and Kigali produced a large increase in importation pressure across the continent (1.4-fold and 1.3-fold, respectively), whereas expansion to Kinshasa, Bujumbura, or Juba produced comparatively limited continental effects. The relative contribution of air and land mobility also varied substantially across scenarios (**Fig. 4c**). Amplification in major transportation hubs markedly increased the contribution of air travel. Ethiopia saw the largest shift, with the contribution of air travel increasing from the current 7% of overall importation risk to 19% under the amplification scenario in Kinshasa. Kenya exhibited a similar pattern, whereas the other countries at highest risk remained dominated by land-mediated importation in all scenarios. All results presented above assumed an epidemic activity of 5 cases per 100,000 persons in the cities considered for the amplification scenarios. Sensitivity analyses using 1 and 10 cases per 100,000 showed that, while the magnitude of amplification varied with epidemic activity, country-level risk rankings remained largely robust (**Supplementary Note 1**). The largest changes occurred for amplification in Lusaka, where increasing epidemic activity shifted Southern African countries towards higher risk.

**Figure 4.**
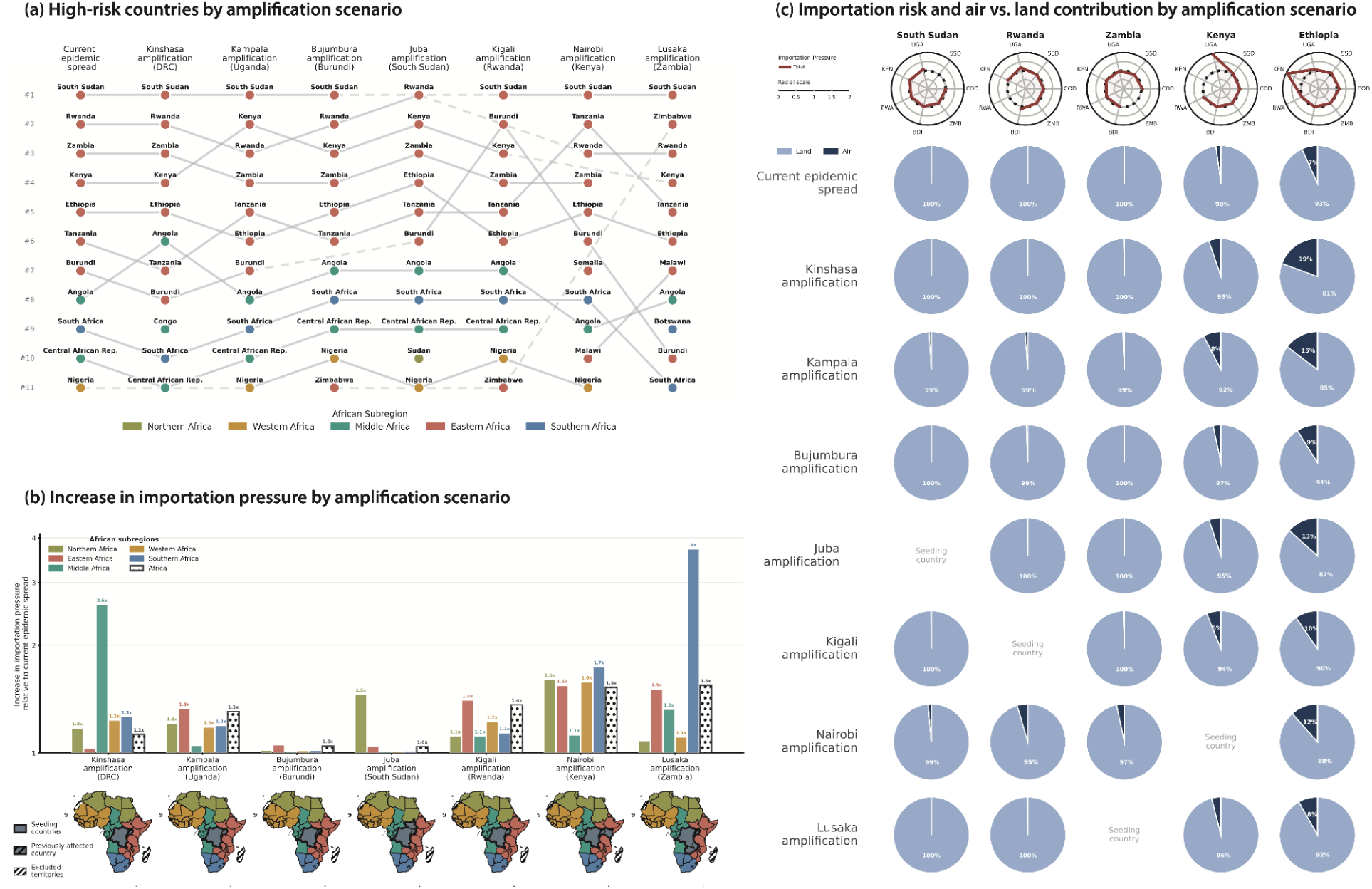
Importation risk and importation pressure to African countries and subregions across different amplification scenarios. Scenarios include Kinshasa, the capital city of the source country (DRC); Kampala, the capital of Uganda, which was previously affected during the outbreak; and the capitals of neighboring countries scoring highest in importation risk. Bujumbura was selected instead of Gitega as it is the economic capital and largest city of Burundi. **(a)** The bump chart shows the change of the rank of high-risk countries (n=11) under the current epidemic spread (first column) and the amplification scenarios. Dots are colored according to subregions. The full ranking is provided in **Supplementary Note 1. (b)** Fold change in importation pressure to African subregions relative to the current epidemic situation under the outbreak expansion scenarios. The dotted bar represents the overall increase for the African continent. The maps show the subregions definitions and the seeding countries (in gray with thicker border) in each scenario. Uganda, being declared Ebola-free, is excluded from the analysis and is shown in grey with black hatching; excluded territories are shown in white with black hatching. **(c)** Top row: Radar plots showing the fold change in importation pressure relative to the current epidemic situation across each amplification scenario for the 5 highest-risk countries. Second to ninth row: Pie charts indicating the within-country shares of overall importation risk attributable to land and air mobility under the current epidemic spread (second row) and under each amplification scenario (from third to ninth row).

## DISCUSSION

We developed an integrated analytical framework combining high-resolution epidemiological and population data with complementary air and land mobility, geographical accessibility, conflict-adjusted travel times, and national preparedness indicators to jointly quantify relative importation risk and national readiness across Africa, in the context of the current outbreak of Bundibugyo Ebola virus disease in eastern DRC. This integrated perspective provides an operational basis for prioritizing preparedness activities, surveillance, and international support during the current outbreak.

Countries at highest risk of Ebola importation differed substantially in their capacity to detect, investigate and respond to imported cases, resulting in a heterogeneous landscape of epidemic vulnerability. Distinct readiness profiles emerged, ranging from countries combining high exposure with strong operational capacity, such as Zambia, Ethiopia and Tanzania, to countries where high importation risk coincides with substantial readiness gaps, including South Sudan, Burundi, and the Central African Republic. South Sudan, the country at highest importation risk, combined intermediate-low emergency preparedness and response with limited healthcare system capacity, raising concerns that imported cases could more readily generate secondary transmission. Looking beyond aggregate readiness scores, the underlying capacity gaps indicate that preparedness priorities should be tailored to country-specific vulnerabilities, spanning human resources, laboratory and infection prevention and control capacity, risk communication, and detection at points of entry. Addressing these gaps would strengthen complementary stages of the response, from early detection and confirmation to containment and outbreak management. This reinforces the importance of continued investments in surveillance, laboratory capacity, emergency preparedness, and cross-border cooperation, consistent with the recent Tabletop Exercise^25^, contingency planning, and preparedness activities coordinated by WHO and Africa CDC. In addition, since 14 July, two fellows from the European Centre for Disease Prevention and Control (ECDC) have also been deployed through the Global Outbreak Alert and Response Network (GOARN) to the WHO country office in Juba, South Sudan, to support surveillance and data analysis activities. At the same time, recent outbreak experience demonstrates that operational preparedness is not solely determined by structural readiness indicators. During the 2024–2025 mpox outbreak, Burundi benefited from the deployment of multidisciplinary African Volunteers Health Corps teams^26^, strengthening local expertise in epidemiology, infection prevention and control, case management, laboratory services, and risk communication. Likewise, Rwanda, a high-risk country with only intermediate readiness scores, successfully contained the 2024 Marburg virus disease outbreak^27^, illustrating how accumulated operational experience and regional coordination can substantially reinforce response capacity. These examples illustrate the broader evolution of epidemic preparedness across Africa.

Repeated responses to Ebola, Marburg virus disease, Sudan virus disease, mpox and cholera, as well as the COVID-19 pandemic experience, have progressively strengthened surveillance, laboratory capacity, emergency operations, and regional coordination, demonstrating the continent’s expanding capacity to control epidemics^28^. The ongoing Bundibugyo outbreak nevertheless highlights that important heterogeneities persist, emphasizing the need to sustain preparedness investments while prioritizing countries where high importation risk coincides with lower readiness.

Our findings are broadly consistent with the current operational prioritization^29^ coordinated by WHO and Africa CDC, with a few important differences. South Sudan, Burundi and Rwanda, currently classified as Priority 1 countries, all emerged among the countries at highest importation risk in our analysis. Ethiopia, Kenya, Tanzania, Zambia, Angola and the Central African Republic, all classified by WHO as Priority 2, were here identified as high-risk destinations. The main exceptions were the Republic of the Congo and Somalia, both classified as Priority 2 but estimated to have low and intermediate-low importation risk in our analysis, respectively. The Republic of the Congo ranked low because the currently affected areas are geographically distant from its borders and therefore contribute little to regional dissemination through land mobility. Somalia, despite being relatively more connected by air than by land, has limited overall connectivity with the affected areas compared with other Priority 2 countries. Conversely, South Africa and Nigeria, although classified as Priority 3 countries by WHO (together with all remaining Member States), were estimated to have a high importation risk despite their greater geographical distance from the outbreak source, reflecting their strong long-range air connectivity. Beyond the identification of countries at highest importation risk, our quantitative estimates of the risk also allow preparedness priorities to be refined further. Botswana, Burkina Faso, Cameroon, Chad, and Ghana emerged as a second tier of priority countries, combining intermediate-high importation risk with important preparedness gaps. Strengthening surveillance and response capacities in these settings could further reduce the risk of regional amplification should the outbreak continue to expand.

The observed geography of importation risk reflects the complementary roles of land and air mobility in regional dissemination. Land mobility dominated the overall pattern of importation risk, generating a strong spatial concentration of risk around the outbreak source that progressively declined with increasing border distance. However, geographical proximity alone was insufficient to identify countries at highest importation risk. As illustrated by the Republic of the Congo, neighbouring countries can differ substantially in exposure depending on the location of the affected areas and their connectivity through regional mobility. For example, under the outbreak amplification scenario in Kinshasa, the Republic of the Congo entered the high-risk group because epidemic expansion into a densely populated area near its borders substantially increased its exposure. Preparedness priorities can therefore rapidly shift as the epidemic expands geographically. The strong dependence on the heterogeneous spatial distribution of epidemic activity and population size may also explain the discrepancies with recent modeling estimates^9^ of cross-border spillover, which classified Rwanda and Burundi as low- and very-low-risk countries, respectively. By explicitly accounting for high-resolution spatial heterogeneity, together with geographically explicit travel times adjusted for conflict, our framework identifies a different pattern of regional dissemination risk. Beyond this strong local component, air transportation remained an important driver of long-range dissemination, elevating the importation risk of countries such as Ethiopia and South Africa, despite their greater border distance from the outbreak source. Regional importation risk cannot therefore be inferred from geographical proximity alone, and – even beyond neighboring countries – analyses based exclusively on air transportation^13^ fail to capture the full geography of regional dissemination. Disentangling air- and land-mediated importation risk is also operationally relevant, because these pathways require different preparedness measures: air-mediated risk points to surveillance at major air transport hubs, whereas land-mediated risk calls for strengthened detection along formal and informal cross-border land mobility corridors.

The outbreak amplification scenarios highlight that preparedness priorities should remain dynamic. The highest-risk countries remained largely unchanged, reinforcing the need for sustained investments in preparedness capacities in these settings. However, epidemic expansion into major regional hubs brought additional countries into the high-risk group. These countries should therefore be considered potential priority targets should the epidemic evolve. Moreover, amplification would substantially increase continental importation pressure, highlighting the importance of anticipating epidemic spread beyond the initial outbreak area. In the absence of licensed vaccines and specific therapeutics against Bundibugyo Ebola virus, strengthening surveillance, laboratory capacity, infection prevention and control, contact tracing, risk communication, and cross-border coordination remains the cornerstone of preparedness^8^.

This study has limitations. First, reliable data on cross-border land mobility in Africa remain scarce. We therefore estimated land mobility from the best available continental dataset, combining multiple mobility sources and deriving land movements as the residual after discounting commercial air travel. Our reconstructed DRC–Uganda flow was of the same order of magnitude as independent estimates reported by McCabe et al^30^, providing an external consistency check for this corridor. However, comparable data were not available for other cross-border flows, preventing validation of the relative magnitude of mobility across corridors. Given this remaining uncertainty, we tested the sensitivity of our results to substantial reductions in land mobility: reductions of 66% and 90% produced only limited changes in country rankings, supporting the robustness of our findings (**Supplementary Note 1**). Second, land-mediated importation risk was estimated assuming an average travel duration of 8h per day, and analyses were restricted to contiguous Africa because comparable data on maritime mobility are not available. Alternative travel durations produced no changes in the estimated geography of importation risk (**Supplementary Note 1**). Third, air-mediated importation risk only considered commercial passenger flights and did not account for humanitarian, military, or charter flights. Fourth, epidemic readiness was assessed using national-level indicators and therefore does not capture subnational heterogeneity in preparedness and response capacity. As a result, countries identified as preparedness priorities may still exhibit substantial geographical variation in their ability to detect and respond to imported cases, depending on where importation occurs. Fifth, both epidemiological and behavioural uncertainties remain. We assumed that infected individuals travelled only during the latent phase, consistent with the rapid onset of symptoms and progressive clinical deterioration characteristic of Ebola virus disease. However, some individuals with early or mild symptoms may still travel before seeking care, particularly over shorter distances, potentially increasing importation risk to more distant destinations. We also assumed homogeneous travel behaviour and exposure risk across travellers, although age, occupation, socioeconomic conditions, purpose of travel, and individual exposure may modify infection risk, as illustrated by the imported case in France involving a medical doctor working in the response^3^. Similarly, geographically heterogeneous under-ascertainment is difficult to quantify. We therefore used the proportion of verified alerts as a proxy for reporting rates by health district, and found the results to be robust to alternative assumptions (**Supplementary Note 1**). Finally, we did not explicitly model behavioural adaptations or the impact of border control measures. While some border restrictions were implemented during the current outbreak, independent mobility data suggested no measurable change in cross-border movements (**Supplementary Note 1**). Moreover, evidence from previous Ebola outbreaks highlights that travel restrictions have limited impact on spatial dissemination while potentially hampering response activities^6^.

Our findings demonstrate that the geography of regional Ebola importation risk in Africa is more complex than proximity to the outbreak alone, emerging from the interplay of local epidemic activity with land and air mobility mediated by geographic accessibility. Our approach provides an operational framework for refining preparedness priorities, updating them as outbreaks evolve and medical countermeasures become available, and anticipating changes before regional dissemination accelerates.

## Data Availability

Proprietary air travel data are commercially available on request from the International Air Transport Association databases (https://www.iata.org/). Importation risk values and readiness score values by countries are available on GitHub at https://github.com/EPIcx-lab/2026-Bundibugyo-Ebola-importation-risk-and-readiness. All other datasets are publicly available at the cited references.

## AUTHORS’ CONTRIBUTION

VC and EV conceived and designed the study. VC supervised the research. FF, FP, BW, MAP collected and analyzed the data. EV, FP, VC developed the analytical framework. FF developed the conflict-adjusted travel-time framework. FF, FP, BW wrote the code and performed the risk analysis. FF, FP, MAP, BW, YB, MF, PM, RLO, VR, YY, CP, EV, VC contributed to the methodology. FF, BW, MAP produced the figures. FF, FP, MAP, BW, YB, MF, PM, RLO, VR, YY, CP, EV, VC interpreted the results. CP, VC wrote the Article. All authors contributed to and approved the final version of the Article.

## ACKNOWLEDGMENTS

This study was partially supported by: the EU Horizon Europe grant ESCAPE (101095619) to F.P., M.A.P., V.C.; the ANRS Maladies Infectieuses Émergentes and France 2030/SGPI grant PReViX (ANRS-24-PEPRMIE-0003) to V.C.; the ANRS Maladies Infectieuses Émergentes ESCAPE project (Start Programme - Fellowships funding scheme - PhD grants 2025 ANRS00850-R-AR1) to F.F., V.C.; the EU Horizon Europe grant SIESTA (101131957) to E.V.; the ANR JCJC grant DiscoReel (ANR-25-CE45-5346-01) to E.V.. F.P., E.V., V.C. acknowledge the Working Group on Pandemic Preparedness of the French Research Action on Modeling Epidemics (FRAME, AC Modélisation des maladies infectieuses) funded by the ANRS Maladies Infectieuses Émergentes.

